# Co-developing a danced version of the Graded Repetitive Arm Supplementary Program (GRASP): a participatory design study with clinicians and people with stroke

**DOI:** 10.1101/2025.09.05.25335089

**Authors:** Marika Demers, Manouchka Louis-Jean, Rovana Keliny, Sylvie Trudelle, Janice J Eng, Diane Leduc, Hanna Pohjola, Annie Rochette, Lucie Beaudry

**Author notes:** Corresponding author: Marika Demers, OT, PhD, School of Rehabilitation, Faculty of Medicine, Université de Montréal, 7077 ave du Parc, Montréal, QC, Canada, H3N 1X7. Disclosure statement: The authors report there are no competing interests to declare.

## Abstract

Self-directed exercise programs empower people with stroke to engage in recovery through daily practice but often present challenges in maintaining long-term adherence. Dance can make rehabilitation exercises more engaging, and help reduce the perception of efforts, due to its inherently enjoyable nature and the use of music. This project aims to co-develop a danced version of the Graded Repetitive Arm Supplementary Program (GRASP) delivered using short video capsules to enhance upper limb motor recovery. Using a participatory research approach, co-development meetings were conducted with researchers and occupational therapists to adapt and refine the GRASP-dance. We beta-tested the GRASP-dance with two people with stroke to refine the program. Feedback from participants and co-development meeting summaries were analyzed using inductive thematic analysis. Two main themes were derived from the co-development meetings: 1) Considerations for the transposition of the GRASP into a danced version, and 2) Challenges and solutions for the development of the GRASP-dance. We adapted the GRASP into a danced version, incorporating the fundamental elements of the GRASP and key considerations at the person, the environment and the dance level. The final program includes 47 video capsules across three difficulty levels along with a weekly virtual group meeting. This work emphasized the need to tailor self-directed exercise, to include a motivating artistic element, and to address key barriers to maximize stroke rehabilitation.

## Introduction

Arm and hand (upper limb) paresis is one of the most common and functionally disabling impairments following stroke.^1–3^ Unfortunately, many stroke survivors experience persistent impairments, particularly in fine motor control and dexterity, which are crucial for functional use of the affected upper limb in everyday activities.^4^ People with stroke often require ongoing rehabilitation services to improve upper limb function after returning home and in the chronic phase of stroke recovery. However, after rehabilitation discharge, they often have few opportunities to continue home exercise and further improve upper limb function.^5^

Self-directed exercise programs delivered at home can empower people with stroke to take an active role in their recovery and encourage daily practice to support long-term recovery.^6,7^ One example of a widely-used, evidence-based self-directed program is the Graded Repetitive Arm Supplementary Program (GRASP).^8^ The GRASP is based on intensive, repetitive and task-oriented practice – key principles of experience-dependent brain plasticity to optimize motor recovery.^9^ The program specifically targets six important skills (strengthening, range of motion, movement repetitions, weight-bearing, trunk control and bilateral movement) and encourages the use of the paretic upper limb in daily activities. The GRASP is typically prescribed for one hour per day using a written manual with pictures and instructions for each exercise to guide in-home practice. Weekly follow-up with a therapist can be done in person (one-on-one or in group)^8^ or remotely (phone^10^ or videoconferencing^11^). Despite favorable results, a major challenge of self-directed programs, such as the GRASP, is maintaining long-term adherence.^5^ This stresses the importance to explore innovative approaches to motivate people to continue rehabilitation efforts after formal rehabilitation has ended.

Clinicians typically provide printed exercise sheets to continue stroke rehabilitation at home, but digital tools can offer more interactive and engaging alternatives. Our team proposes to adapt the GRASP into a danced version delivered using short video capsules for self-directed practice supported by virtual weekly group meetings. We specifically selected dance, as it inherently integrates fundamental motor learning principles such as task-specific practice, sensory feedback, and social interaction.^12,13^ Dance incorporates music and rhythm, which may make exercises more engaging.^14^ The choreography, repertoire, and quality of the movements distinguish dance from exercise.^15^ Recent evidence suggest that clinicians, people with stroke and their loved ones perceived positively dance interventions to foster motivation, self-efficacy, and social interactions.^16^ Dance can also reduce the perception of efforts in people with stroke.^17^ While dance may be promising, challenges in adapting a structured exercise program into a danced version, such as movement modifications for different abilities and a balance between structure and creative expression should be considered.

This paper aims to report on the co-development process of a danced version of the GRASP to enhance upper limb motor recovery in people with stroke.

## Material and methods

### Study design

This study used a participatory research approach in five phases to develop a danced version of the GRASP (GRASP-dance): 1) Preparatory, 2) Precision, 3) Structuring, 4) Development and 5) Improvement.^18,19^ This approach proposes an iterative process in which the development and testing are concomitant and interdependent stages, evolving according to the reflections and data collected. This study is a first step in a larger project aiming at testing the feasibility and the efficacy of the GRASP-dance. Only the development of the GRASP-dance is presented here to provide insight into how evidence-based therapy can be adapted through creative movement to guide the adaptation of other programs or populations.

### Participants

The co-development team comprised of researchers with expertise in dance (LB, HP), digital pedagogy (DL) and stroke rehabilitation (i.e., physical and occupational therapists; JE, HP, AR, MD), and occupational therapists (n=4) working in stroke rehabilitation, with at least one year of clinical experience in stroke rehabilitation and familiar with the GRASP. Occupational therapists were recruited by email invitations from three partner sites in the greater Montreal. Two patient-partners were also recruited to test a preliminary version of the GRASP-dance using the same inclusion criteria as the virtual GRASP.^11^ Specifically, community-dwelling chronic stroke survivors with upper limb motor impairments were included. Participants with significant pain preventing movement of the upper limb were excluded. This project was approved by the Comité d’éthique de la recherche en réadaptation et en déficience physique (MP-50-2023-1639) and the Comité institutionnel d’éthique de la recherche avec des êtres humains of the Université du Québec à Montréal (2023-5102). All participants were informed of the procedures and the nature of their participation and provided written informed consent.

### Graded Repetitive Arm Supplementary Program

The GRASP is a self-directed program designed to improve upper limb strength, coordination and use in daily activities through task-specific, intensive and repetitive practice.^8,10^ The program includes 35 exercises divided into 5 sections: 1) Stretching, 2) Arm strengthening, 3) Hand strengthening, 4) Coordination and 5) Hand skills. People with stroke are asked to complete one hour of prescribed exercises per day and are monitored weekly throughout the course of the program. For each participant, a therapist prescribes the program with set repetitions to meet the need of each person with stroke, teaches the program and monitors progress weekly. The weekly monitoring can be done in person,^8^ remotely over the phone^10^ or in a group session through a videoconferencing platform.^11^ During weekly monitoring, the program is adjusted by varying the number of repetitions and behavioral strategies are offered to promote greater use of the affected upper limb. An exercise book with descriptions and pictures of each exercise is offered to guide self-directed practice. The required equipment to complete the GRASP includes inexpensive and everyday items such as a towel, a ball, blocks and paper clips.

### Procedures

Throughout the five co-development phases, we conducted co-development meetings with the researchers and with the occupational therapists. We iteratively developed and refined the GRASP-dance. We produced alpha-versions of short video clips displaying dance routines combining GRASP exercises. We showed the video clips during the co-development meetings to gather feedback and improve the program. Each co-development meeting was aligned with a phase of development and was articulated over a working theme. Table 1 provides an overview of the working themes for each phase. The co-development meetings were done remotely using the videoconferencing platform (Zoom telecommunications inc., San Jose, CA), with separate meetings for the researchers and the occupational therapists.

**Table 1.** Working theme for each phase of co-development.

| Phase | Stakeholders | Working themes | GRASP sections |
| --- | --- | --- | --- |
| Preparatory | Researchers | - Fundamental elements of the GRASP<br>- Potential challenges for dance transposition<br>- Strategies to adapt the GRASP into a danced version | Program structure |
| Refinement | Researchers<br>Clinicians | - Structure of GRASP-dance<br>- Movement proposals<br>- Music proposals | Program structure<br>Sections 1-3 |
| Structuring | Researchers<br>Clinicians | - Required equipment and incorporation of props<br>- Transposition of the movements into a danced version<br>- Themes and metaphors for routines | Sections 2-4 |
| Development | Researchers<br>Clinicians | - Strategies to achieve repetition targets and promote interactivity and engagement<br>- Gradation of the difficulty level<br>- User and instructor manuals | Sections 1-5 |
| Beta-testing with patient-partners |  |  |  |
| Improvement | Researchers<br>Clinicians | - Challenges experienced during beta-testing and potential solutions<br>- Video production<br>- Considerations for a clinical trial | Sections 1-5 |

### Preparatory phase

The co-development began with a 60-min meeting with the research team to identify the format of the GRASP-dance, the key elements from the GRASP to maintain in the adaptation and potential issues in transposing the GRASP into the GRASP-dance. The research team elaborated an evaluation grid describing the fundamental elements of the GRASP and the targeted skills for each exercise to guide subsequent phases of development. Movement sequences were developed for the Stretching, Arm and Hand strengthening sections.

### Phases 1 to 3 - Refinement, Structuring and Development

These phases aimed to refine, structure and develop the GRASP-dance. Preliminary versions of the program, including beta version of the video clips, music inspirations, and drafts of the user and instructor manuals, were presented during the co-development meetings. Ad hoc individual discussions were held with a subset of the research team to brainstorm or refine specific components of the program, such as sequences of movements combining different exercises, themes and metaphors for the dance routines and strategies to encourage interactions.

### Beta-testing with people with stroke

We beta-tested a preliminary version of the GRASP-dance with the two patient-partners. We purposefully recruited one man and one woman with different severity of sensorimotor impairments. The program was tested in a single session with a dance instructor delivering the program with the assistance of an occupational therapist. One session was offered remotely using the Zoom platform (Zoom Video Communications, San José, CA), whereas one session was done in hybrid (30 min over Zoom on a tablet and 30 min in person) to understand the different environments and provide hands-on assistance for the participant with more severe motor impairments. Before the start of the beta-testing, the occupational therapist assessed the sensorimotor impairments severity to identify the version of the routines to be used. During the beta-testing, the occupational therapist made suggestions of adaptations when difficulty arose or provided additional instructions. Fatigue and pain were assessed on a visual analogue scale pre- and post-testing session. Both sessions were video recorded to identify difficulties encountered and spontaneous feedback from participants and/or the occupational therapist. After the testing, we conducted a short semi-structured interview with each participant on the experience with the GRASP-dance, the perceived relevance, the difficulties experienced and potential changes to make to the program. We also gathered the experiences of the therapist delivering the intervention through a semi-structured interview. The semi-structured interviews with the patient-partners and the therapist were included in the analysis.

### Phase 4 - Improvement

A summary of the data collected during the beta-testing was presented at the beginning of the co-development meeting to identify modifications to be made to the GRASP-dance and find solutions to the challenges encountered. Following the modification of GRASP-dance, we held a final co-development meeting to prepare the final production of the video capsules and user and instructor manuals.

### Data analysis

A detailed summary of each co-development meeting was produced to capture all the ideas expressed. The semi-structured interviews were transcribed verbatim. Two members of the research team (MLJ and RK) independently analyzed the summaries of the co-development meetings and the transcribed verbatim using an inductive approach to thematic analysis based on Braun and Clark framework.^20^ A detailed codebook with definitions for each code was developed. A third researcher (MD) was involved in guiding reflections on codes and themes, and in resolving disagreements between both coders, while a fourth researcher (LB) was involved to validate the coding and themes. The analysis was carried out using QDAMiner software (Provalis Research, Montreal, Canada). The data were triangulated across coders and between the perspectives of patient-partners and clinicians during the development session. Regular analytical meetings we used to compare interpretations and verify accuracy. Throughout the co-development and data analysis, we also used a detailed logbook to keep track of decisions and modifications made and reflections. Descriptive statistics were used for the demographic, fatigue and pain data.

### Reflexivity

The qualitative data analysis was performed by two graduate students in occupational therapy (MLJ and RK), under the supervision of an assistant professor in occupational therapy (all women). We purposefully selected two people who were not involved in the co-development process to perform the analysis to offer an independent interpretation of the data. Both graduate students had prior experience in stroke rehabilitation but only received basic training on the GRASP before starting the data analysis. MD is an assistant professor with experience in qualitative research with people with stroke, and is familiar with the GRASP program. LB is an associate professor in dance, with experience with adapted dance for people with stroke and qualitative research. Both oversaw the study design and implementation. Critical self-examination and discussions took place between members to recognize and reflect on the influence of different life experiences on the data analysis.^21^ We acknowledged the co-constructed nature of qualitative knowledge and remained attentive to how our own assumptions, professional backgrounds, and positionalities shaped the research process and interpretation of participants’ contributions. A reflexive journal was also used to support the confirmability and transparency of the qualitative findings and provide reflections on any assumptions and interpersonal dynamics.

## Results

Four occupational therapists and six interdisciplinary researchers with expertise in stroke rehabilitation (n=4), dance (n=2) and digital pedagogy (n=1), including the creator of the GRASP (JE), participated to the co-development of the GRASP-dance. The co-development team members were all women. Two people with stroke (1 man/1 woman, severe/mild motor impairments, respectively) participated in the beta-testing (see Table 2 for detailed participant characteristics). A caregiver was not present in the beta-testing sessions.

**Table 2.**
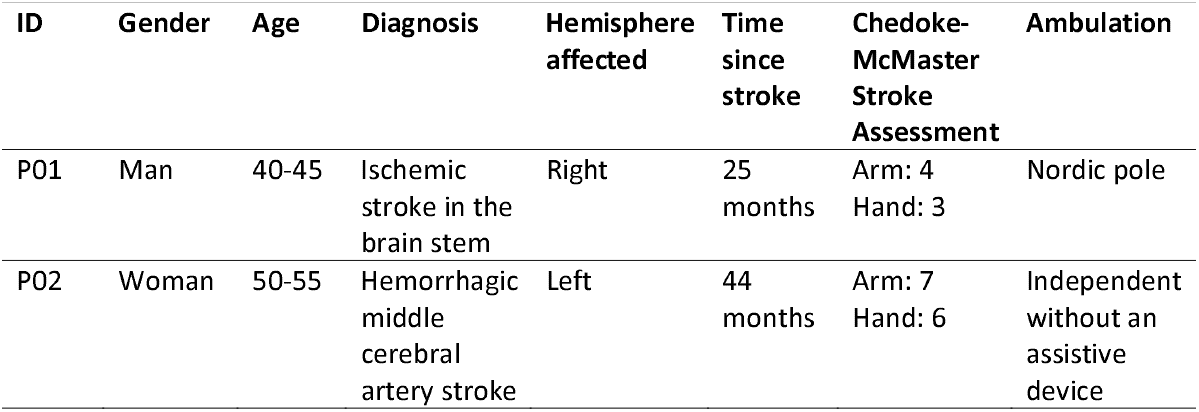
Participant characteristics.

| **ID** | **Gender** | **Age** | **Diagnosis** | **Hemisphere affected** | **Time since stroke** | **Chedoke-McMaster Stroke Assessment** | **Ambulation** |
| --- | --- | --- | --- | --- | --- | --- | --- |
| P01 | Man | 40-45 | Ischemic stroke in the brain stem | Right | 25 months | Arm: 4<br>Hand: 3 | Nordic pole |
| P02 | Woman | 50-55 | Hemorrhagic middle cerebral artery stroke | Left | 44 months | Arm: 7<br>Hand: 6 | Independent without an assistive device |

The results section presents the main themes from the co-development meetings and beta-testing, and a summary of the final GRASP-dance program version. The two main themes are: 1) Considerations for the transposition of the GRASP into a danced version, and 2) Challenges and solutions for the development of the GRASP-dance. The challenges and solutions were organized according to the person, environment and dance.

### Theme 1. Considerations for the transposition of the GRASP into a danced version

This theme refers to the fundamental elements of the original GRASP to retain, and the structural, and conceptual adaptations required to translate this program into a danced version. Researchers and clinicians emphasized that core components of the GRASP and the seven ‘must-do’ parts, such as the exercise progression and the generalization of skills towards functional tasks, were crucial to include in the GRASP-dance (Figure 1 displays the key components of GRASP maintained in the adaptation with an example of a routine). The virtual weekly group meetings were judged important to discuss the challenges encountered by people with stroke, review the routines, make suggestion of adaptations and foster accountability. The logbook was also identified as an essential tool for monitoring participants’ progress and practices. The co-development team recognized that target repetitions, dosage, and intensity should be carefully calibrated in the GRASP-dance to align with those of the original. As a strategy to encourage the transfer of acquired skills to everyday activities, it was proposed to link each routine to a list of functional activities and review those during weekly meetings.

**Figure 1.**
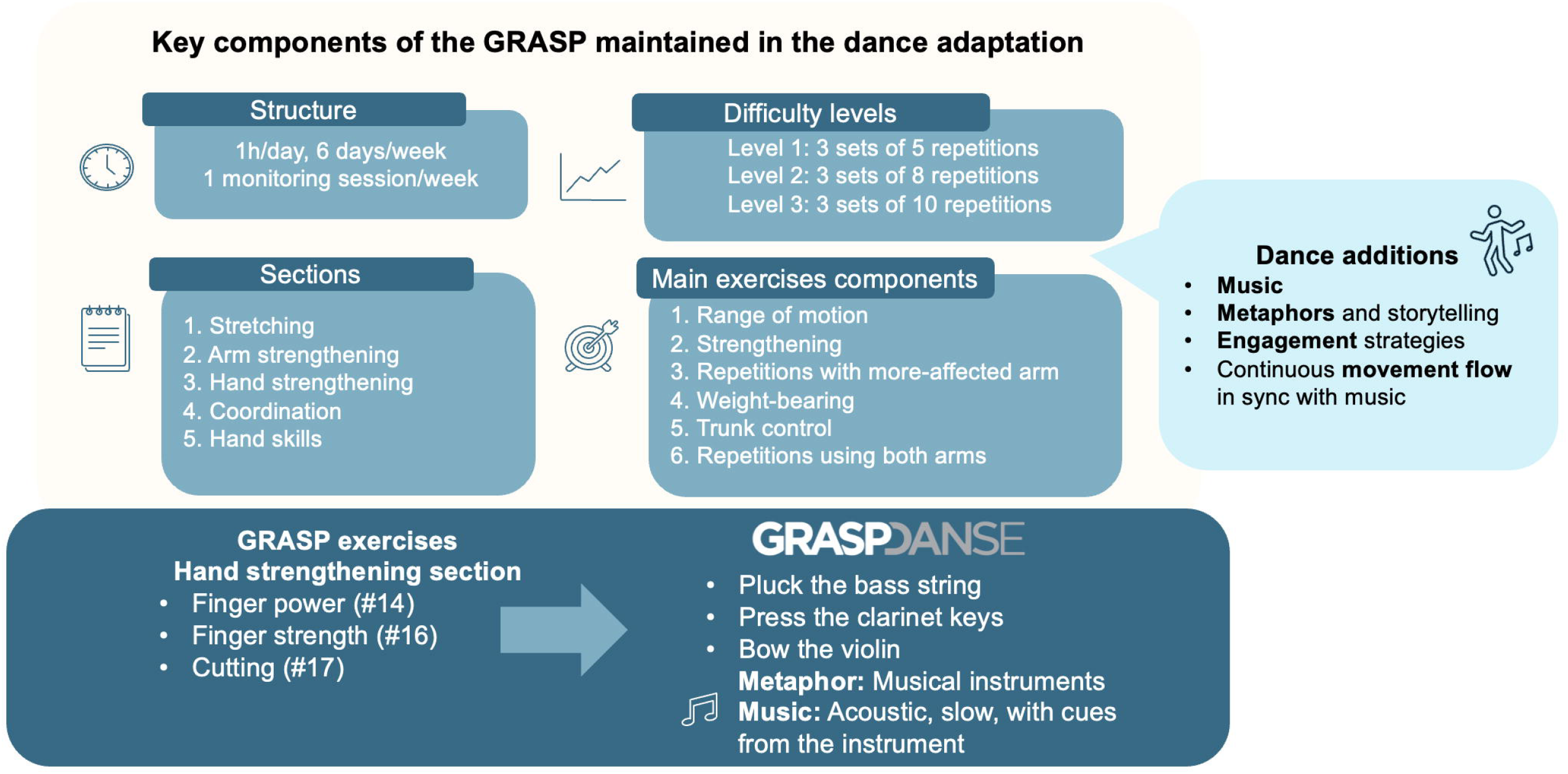
Key components of the Graded Repetitive Arm Supplementary Program (GRASP) maintained in dance adaptation.

The co-development team identified key features of the GRASP-dance, such as the use of music and the playful nature of dance, which can reduce the perception of physical effort and pain. The use of metaphors and story-inspired sequences was identified as key to maintain the artistic component of dance. A combination of several exercises in a single routine was preferred over performing each exercise in succession. Nevertheless, participants suggested organizing the exercises into groups corresponding to each of the five sections of the GRASP (i.e., stretching, arm and hand strengthening, coordination, hand skills) to guide clinicians in selecting the right routines. The manipulation of objects rather than mimicking the movements without an object was considered essential. However, it was suggested to reduce the number of props to a few affordable, versatile, everyday objects.

### Theme 2. Challenges and solutions for the development of the GRASP-dance

This theme encompasses potential barriers to the transposition of the GRASP into a danced version and proposed strategies to overcome each barrier (Table 3 displays a summary of the main challenges and solutions identified with representative quotes).

**Table 3.**
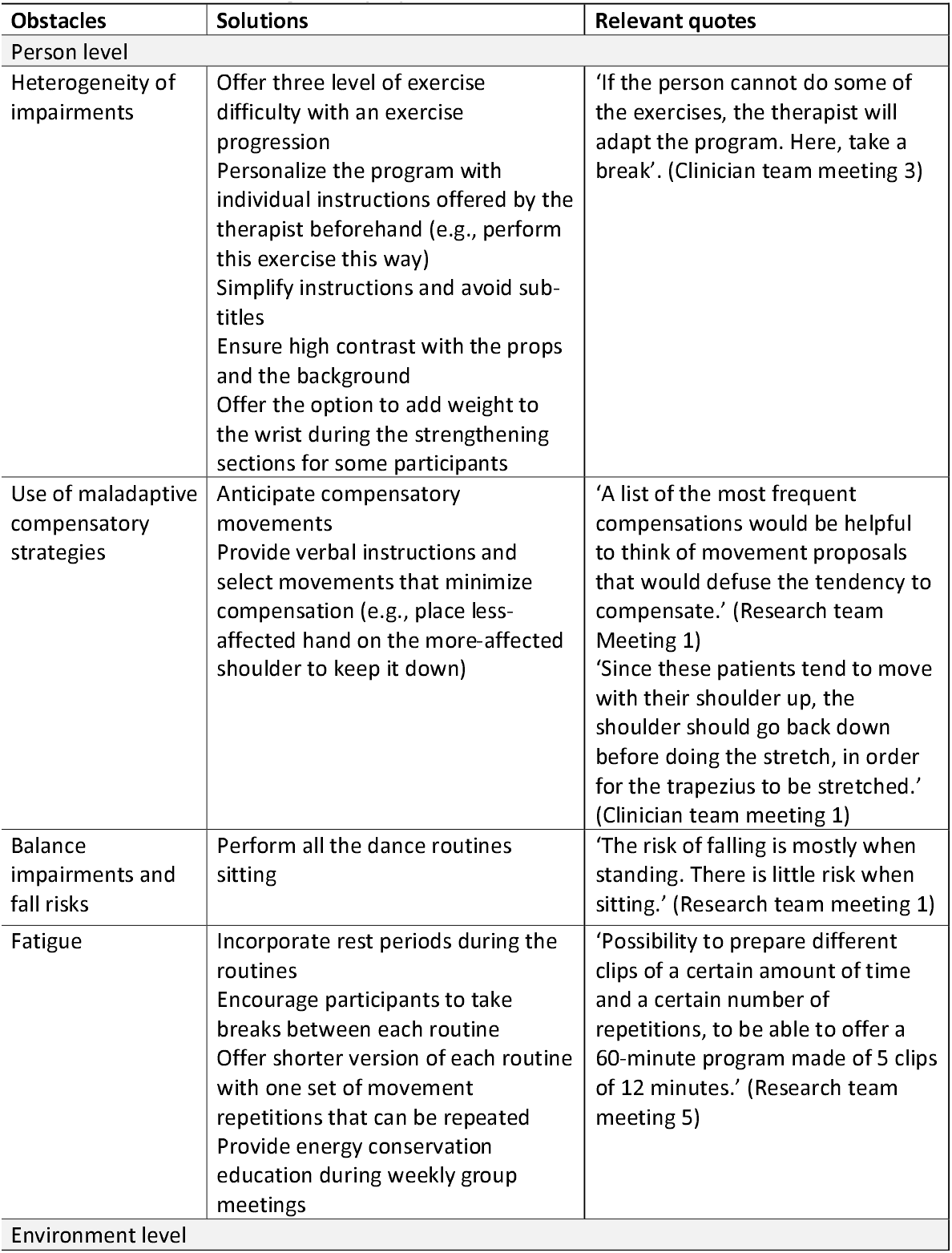
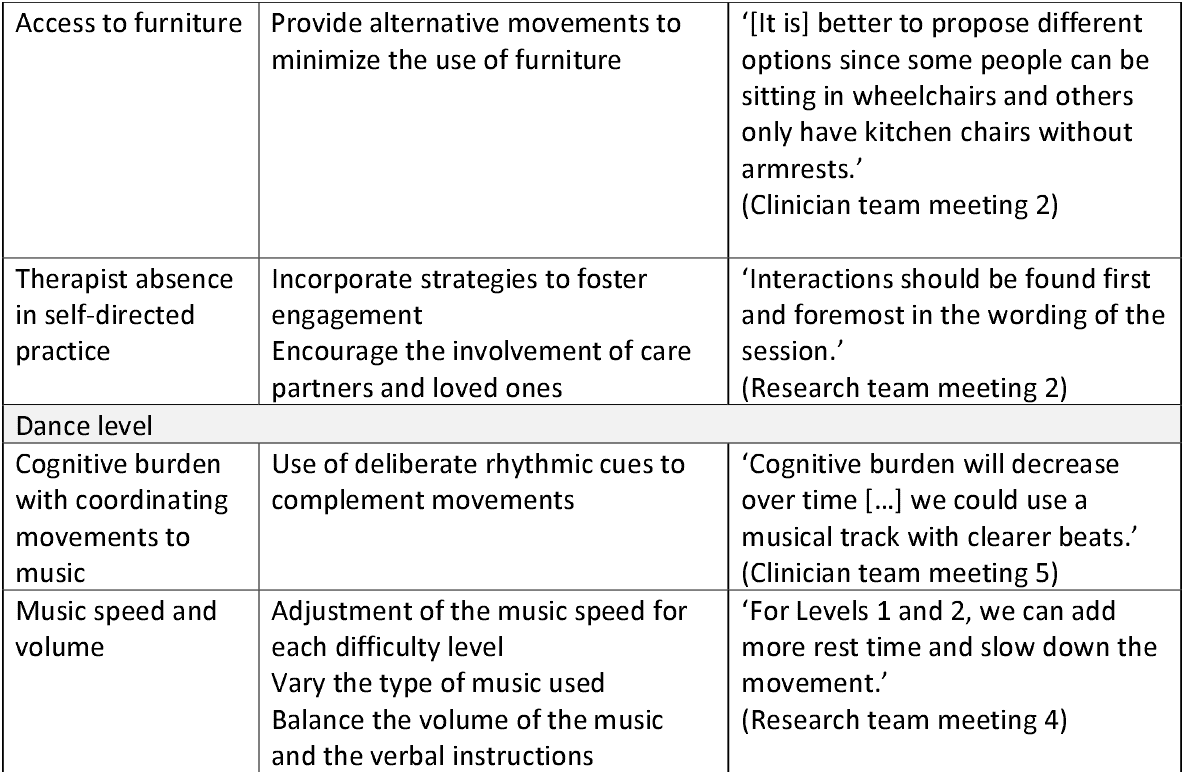
Overview of challenges and proposed solutions.

### 2.1 Person

The heterogeneity of post-stroke impairments was identified as the main challenge in creating video capsules for self-directed practice. Participants stressed the importance to especially consider cognitive, language and visual perceptual impairments, in addition to the wide range of sensorimotor impairments common after stroke. People with stroke may adopt maladaptive compensatory strategies during self-directed practice, such as excessive shoulder elevation or trunk flexion. One solution was to anticipate compensatory movements and to provide verbal instructions and select movements that minimize compensation. It was agreed that all routines should also be performed seated to reduce the risk of falls or injury. To minimize fatigue, suggestions were made to divide the GRASP-dance into smaller sections, incorporate rest periods during the routines and offer a short version of the routines with one set of movement repetitions that can be repeated.

### 2.2 Environment

The GRASP requires the use of furniture, such as a chair with armrests. Participants suggested alternative exercises to allow people with stroke to participate regardless of the furniture at home. Due to the self-directed nature of the program, strategies to foster engagement were identified, such as having the instructor in the video capsules speak directly to participants and choosing deliberately the wording of the instructions and cues during the routines. Care partners and loved ones were also considered important to encourage adherence and accountability (e.g.: set-up equipment, complete the program’s logbook, provide encouragements).

### 2.3 Dance

The GRASP involves mostly unimanual exercises with the more-affected side. In the danced version, it was suggested to incorporate more bimanual or bilateral movements but maintain the emphasis on the more-affected side. While music and its rhythmic cues may support movement execution and make daily practice more engaging than typical self-directed exercise programs, participants voiced concerns that coordinating movements to music may increase the cognitive burden on people with stroke. The use of deliberate rhythmic cues was suggested to complement movements and highlight elements, such as a new set of movement repetition or a change of exercise. Another consideration was the speed of the music; a rapid tempo may limit one’s ability to follow the routine, but slow tempo may not be as engaging. The importance to adjust the volume of the music to the verbal instructions was also raised to help people with stroke to follow the instructions.

### Beta-testing

Both participants reported high levels of satisfaction with the GRASP-dance. They also perceived benefits of a single session for their upper limb. There was no pain in the upper limb before and after the test (mean pre: 0, post: 0). Perceived fatigue increased on average from 1.5/10 before the testing session to 4.5/10 at the end. The participants generally felt that the session was valuable, and that the intervention was relevant to their rehabilitation. They also both spontaneously reported perceived benefits for their upper limb, despite having done a single session.

> P01 ‘It makes my arm more flexible […] I feel like my arm is constantly improving when moving. I’m just not use to using it.’ ‘I really enjoyed the music. Slower music made it easier to understand how to perform the movement. If the rhythm was too fast, it increased my spasticity.’
>
> P02 ‘The whole idea is excellent […] The rhythm helps me to relax.’ ‘I liked the variety of props.’ ‘I liked the metaphors, that’s the thing that helped me focus more, because I was listening. I could close my eyes.’ ‘We all know how to dance to some extent, but often we’re afraid. Here, on the other hand, you feel free. You can do the movements more or less the way you feel good. That part is very important because we feel judged too often. […] There’s more flow.’

Challenges were also experienced during the testing session. The 60-minute session felt long for both participants. A suggestion was to break down the session into two shorter 30-minute sessions per day. One participant reported that musical cues could be enhanced to minimize the verbal instructions. For the remote session, the position of the tablet to properly see the video routines while the whole upper body remained in view required multiple adjustments. Since a preliminary version of the videos was showing a participant with right-sided hemiparesis, P02 had to transpose the movements to her more-affected side, creating an additional cognitive load. It was suggested to mirror the videos for participants with left-sided hemiparesis.

### GRASP-dance

We developed 47 video capsules across 3 difficulty levels, a user and an instructor manuals. Each capsule was mirrored to show right- and left-sided hemiparesis (94 capsules in total). Each capsule targeted a specific section of the GRASP and was often associated with a metaphor (e.g. musical instruments, cloud, wings, etc.). The number of movement repetition for each exercise matched the GRASP. A shorter version of each routine with one set of movement repetition was also created to be repeated for participants with lower endurance. The capsules can be combined to reach the target of 60 minutes of daily exercise (average duration: 7.0 minutes, range: 3.3 and 12.4 minutes). Eight props were used to complete all the routines: a loofah sponge, five foam circles (9.5 cm in diameter), five silicone squares (10 cm), a scarf (48 by 148 cm length), a wooden stick (30.5 cm in length, 0.8 cm in diameter), four cubes (three rigid cubes: 5 cm, one foam cube: 10 cm), a ball (7 cm in diameter) and a piece of ribbon (3.5 cm width, 100 cm length). A typical week of the GRASP-dance was structured around an hour of self-directed practice per day guided by video capsules and a weekly remote monitoring session done in a closed group of up to five people, for two months in total.

## Discussion

Using a participatory research approach, we adapted the GRASP into a danced version, incorporating the fundamental elements of the GRASP and key considerations at the person, the environment and the dance level. The final GRASP-dance includes video capsules across three difficulty levels to be combined to achieve an hour daily of self-directed practice, along with a weekly virtual group meeting. This work highlighted the importance to individualize self-directed exercise programs, integrate a motivating artistic component and overcome physical, cognitive and environmental barriers to maximize the benefits of stroke rehabilitation. GRASP-dance has the advantage here of focusing on arm and hand movements, thus avoiding the problem of space and freedom of movement that can be associated with an online dance program.^22^

The GRASP incorporates key principles of experience-dependent neuroplasticity, such as high intensity, repetitive and salient practice.^9^ During the development of the GRASP-dance, a great emphasis was placed on maintaining the key active ingredients of the GRASP (e.g., high repetition, gradation of exercises and generalization of skills to functional tasks), known to optimize functional recovery.^8,10,11^ A delicate balance between structure and creative expression was needed in the adaptation into the GRASP-dance to remain true to the original program. Unlike many dance-based interventions,^12^ the GRASP-dance is very structured, with limited improvisation to meet the repetition targets. Nevertheless, we incorporated various artistic elements—such as storytelling, metaphor, continuous movement flow synchronized with music, and a strong emphasis on timing and rhythm—to ensure the intervention retained its identity as a dance-based practice rather than merely exercises set to music. The GRASP-dance is also well-aligned with four key elements of dance-based interventions hypothesized to impact health and wellbeing in older adults: 1) Balance challenges; 2) Motor-skill learning challenges; 3) Cognitive challenges; and 4) Creative and artistic opportunities.^23^ While the danced version of the GRASP integrates the benefits of using dance, it remains grounded in the structure and key elements of the original program.

The heterogeneity of post-stroke impairments was an important consideration in the development of the GRASP-dance. It is well-recognized that a one-size-fits-all model in stroke rehabilitation is often inadequate for addressing the diverse needs of people with stroke, reinforcing the growing shift toward precision rehabilitation.^24,25^ The GRASP-dance offers routines at three difficulty levels with movement modifications for different abilities (i.e., movement complexity, speed, precision, number of movement repetition), allowing participants to progress through the levels as they improve. While a routine may not match the exact needs of a participant, a therapist can suggest additional adaptations during weekly follow-up meetings. This is consistent with the literature on the importance of providing the right level of challenge to maximize motor learning and neuroplasticity.^26,27^ It is also a strategy to promote safety of self-directed practice to ensure that each capsule is tailored to the individual’s abilities. Several measures were implemented to ensure participants’ safety at home, including conducting the program in a seated position, confirming they had a stable, non-slip chair, incorporating short rest periods in each session, and encouraging the involvement of loved ones. By incorporating modular choreography and an accessible digital format, the GRASP-dance offers an inclusive solution to people with stroke with varying abilities.

Adherence to self-directed practice can be a challenge for people with stroke.^5^ We incorporated multiple strategies to foster engagement: instructions provided directly to the camera, a behavioral contract, a logbook and weekly group meetings. Many strategies were already implemented in the GRASP delivered remotely and known to be effective.^11^ The results from a recent systematic review on self-directed upper limb programs suggest that a strong focus on the practice of tasks directly associated with daily activities may foster higher compliance and lower attrition.^6^

## Limitations

The participating occupational therapists all worked in urban, university-affiliated rehabilitation centers. While their experience was valuable to help shape the GRASP-dance, assumptions about access to technology or equipment that might not hold true in rural or under-resourced settings. It may also limit the generalizability of the findings to stroke care in more rural areas. We beta-tested the GRASP-dance in a single session with two participants. This represents a small sample size that is far from representative of the population under study. Challenges with long-term adherence to the GRASP-dance and with the telerehabilitation platform remain to be tested with a large, diverse sample. Despite these limitations, the co-development approach will ensure that the program is aligned with the needs of people with lived experience of stroke and clinicians. This co-development model could also be used in other contexts, such as the adaptation of other programs or for other populations with movement impairments.

A danced version of the GRASP program was developed in close collaboration with key stakeholders with the aim to optimize upper limb functional recovery. The GRASP-dance maintains the core components of the GRASP, while also incorporating musical and artistic components of dance, as well as original movements. The adaptation of a structured exercise-based program into a dance version requires careful consideration to meet the heterogenous needs of each person with stroke and accounting for differences in the physical and social home environment of each participant. A structured dance format was selected to meet the number of movement repetitions for each level of difficulty. The GRASP-dance included the manipulation of real objects to provide sensory input and incorporated metaphor and storytelling, deliberate rhythmic cues, and movements minimizing maladaptive compensatory movements. As a next step, the feasibility of delivering the GRASP-dance remotely over two months should be established. The perspective of clinicians in delivering this type of creative exercise program should be explored. The effectiveness of the GRASP-dance also remains to be tested in a large, adequately powered non-inferiority trial.

## Data Availability

All data produced in the present study are available upon reasonable request to the authors.

## References

1. Dalton EJ, Jamwal R, Augoustakis L, et al. Prevalence of Arm Weakness, Pre-Stroke Outcomes and Other Post-Stroke Impairments Using Routinely Collected Clinical Data on an Acute Stroke Unit. Neurorehabil Neural Repair. 2024;38(2):148–160. doi:10.1177/15459683241229676

2. Nakayama H, Jorgensen HS, Raaschou HO, Olsen TS. Recovery of upper extremity function in stroke patients: the Copenhagen Stroke Study. Archives of physical medicine and rehabilitation. 1994;75:394–398.

3. Simpson LA, Hayward KS, McPeake M, Field TS, Eng JJ. Challenges of Estimating Accurate Prevalence of Arm Weakness Early After Stroke. Neurorehabil Neural Repair. 2021;35(10):871–879. doi:10.1177/15459683211028240

4. Chen SY, Winstein CJ. A systematic review of voluntary arm recovery in hemiparetic stroke: Critical predictors for meaningful outcomes using the international classification of functioning, disability, and health. Journal of Neurologic Physical Therapy. 2009;33(1):2–13. doi:10.1097/NPT.0b013e318198a010

5. Miller KK, Porter RE, DeBaun-Sprague E, Van Puymbroeck M, Schmid AA. Exercise after Stroke: Patient Adherence and Beliefs after Discharge from Rehabilitation. Topics in Stroke Rehabilitation. 2017;24(2):142–148. doi:10.1080/10749357.2016.1200292

6. Da-Silva RH, Moore SA, Price CI. Self-directed therapy programmes for arm rehabilitation after stroke: a systematic review. Clin Rehabil. 2018;32(8):1022–1036. doi:10.1177/0269215518775170

7. Fryer CE, Luker JA, McDonnell MN, Hillier SL. Self management programmes for quality of life in people with stroke. Cochrane Database of Systematic Reviews. 2016;(8). doi:10.1002/14651858.CD010442.pub2

8. Harris J, Eng JJ, Miller W, Dawson A. A Self-Administered Graded Repetitive Arm Supplementary Program (GRASP) Improves Arm Function During Inpatient Stroke Rehabilitation. Stroke. 2009;40(6):2123–2128. doi:10.1161/STROKEAHA.108.544585

9. Kleim JA, Jones TA. Principles of experience-dependent neural plasticity: implications for rehabilitation after brain damage. Journal of Speech, Language, and Hearing Research. 2008;51(1):S225–S239.

10. Simpson LA, Eng JJ, Chan M. H-GRASP: the feasibility of an upper limb home exercise program monitored by phone for individuals post stroke. Disability and Rehabilitation. 2017;39(9):874–882. doi:10.3109/09638288.2016.1162853

11. Yang C ling, Waterson S, Eng JJ. Implementation and Evaluation of the Virtual Graded Repetitive Arm Supplementary Program (GRASP) for Individuals With Stroke During the COVID-19 Pandemic and Beyond. Phys Ther. 2021;101(6):pzab083. doi:10.1093/ptj/pzab083

12. Kipnis D, Kruusamäe H, King M, Schreier AR, Quinn L, Shih HJS. Dance interventions for individuals post-stroke - a scoping review. Top Stroke Rehabil. 2023;30(8):768–785. doi:10.1080/10749357.2022.2107469

13. Teixeira-Machado L, Arida RM, de Jesus Mari J. Dance for neuroplasticity: A descriptive systematic review. Neuroscience & Biobehavioral Reviews. 2019;96:232–240. doi:10.1016/j.neubiorev.2018.12.010

14. Kattenstroth JC, Kalisch T, Holt S, Tegenthoff M, Dinse HR. Six months of dance intervention enhances postural, sensorimotor, and cognitive performance in elderly without affecting cardio-respiratory functions. Frontiers in Aging Neuroscience. 2013;5(FEB):1–16. doi:10.3389/fnagi.2013.00005

15. Elmslie C, McCallion L, Vaughan-Graham J, Patterson KK. Dance harnesses humanity in exercise: perceptions of dance following an adapted dance program for people with chronic stroke. Arts & Health. Published online February 6, 2025:1–17. doi:10.1080/17533015.2025.2461687

16. Beaudry L, Fortin S, Rochette A. Adapted dance used in subacute rehabilitation post-stroke: impacts perceived by patients, relatives and rehabilitation therapists. Disability and Rehabilitation. 2020;42(21):2997–3006. doi:10.1080/09638288.2019.1581845

17. Beaudry L, Rochette A, Fortin S. Use of Adapted Dance to Intensify Subacute Rehabilitation Post-Stroke: A Qualitative Study on the Participation Experience and Active Participation Time. ALTERN THER HEALTH MED. 2022;28(7):40–51.

18. Bergeron L, Rousseau N. La Recherche-Développement En Contextes Éducatifs. Presses de l’Université du Québec; 2021. Accessed February 18, 2025. https://www.entrepotnumerique.com/p/114762?f=pdf

19. Loiselle J, Harvey S. La recherche développement en éducationlZ:fondements, apports et limites. Recherches qualitatives. 2007;27(1):40. doi:10.7202/1085356ar

20. Braun V, Clarke V. Thematic Analysis: A Practical Guide. SAGE Publications; 2021.

21. Dodgson JE. Reflexivity in Qualitative Research. J Hum Lact. 2019;35(2):220–222. doi:10.1177/0890334419830990

22. Walton L, Domellöf ME, Neely AS. On-line vs. On-site Dance for People with Parkinson’s Disease: An Evaluation Study. NJACH. 2022;4(2):1–13. doi:10.18261/njach.4.2.4

23. Waugh M, Jr GY, Casale C, Balaban R, Cross ES, Merom D. The use of dance to improve the health and wellbeing of older adults: A global scoping review of research trials. PLOS ONE. 2024;19(10):e0311889. doi:10.1371/journal.pone.0311889

24. French MA, Roemmich RT, Daley K, et al. Precision rehabilitation: optimizing function, adding value to health care. Archives of Physical Medicine and Rehabilitation. 2022;103(6):1233–1239. doi:10.1016/j.apmr.2022.01.154

25. Lin DJ, Stein J. Stepping Closer to Precision Rehabilitation. JAMA Neurology. 2023;80(4):339–341. doi:10.1001/jamaneurol.2023.0044

26. Guadagnoll MA, Lee TD. Challenge Point: A Framework for Conceptualizing the Effects of Various Practice Conditions in Motor Learning. Journal of Motor Behavior. 2004;36(2):212–224. doi:10.3200/JMBR.36.2.212-224

27. Plautz EJ, Milliken GW, Nudo RJ. Effects of repetitive motor training on movement representations in adult squirrel monkeys: Role of use versus learning. Neurobiology of Learning and Memory. Published online 2000. doi:10.1006/nlme.1999.3934

